# Indocyanine Green Lymphography Imaging of Normal Lymphatic Drainage in the Lower Limbs

**DOI:** 10.1101/2025.10.06.25337158

**Authors:** Mike Mills, Malou van Zanten, Greta Brezgyte, Bernard Ho, Julian Pearce, Manan Shelton, Peter Mortimer, Hiroo Suami, Kristiana Gordon, Pia Ostergaard

## Abstract

**Objectives:** Indocyanine Green Lymphography (ICGL) has emerged as a potentially powerful tool for the study of the superficial lymphatic system and to support the diagnosis of lymphoedema. However, detailed descriptions of ICGL findings in healthy individuals are limited. In this study, we imaged a series of healthy participants using ICGL, attempting to establish what represents normal presentation.

**Methods:** Sixteen healthy individuals aged 20-55 years were recruited to undergo lower limb ICGL. Diluted 0.1mL injections of ICG were administered intradermally to five sites: 1^st^ and 4^th^ web spaces, lateral midfoot and lateral and medial rear-foot (heel); locations considered optimal for lymphatic vessel access. Outcome measures included: i) the drainage routes of contractile lymphatic collectors observed, ii) the number of lymphatic vessels crossing the anterior ankle and iii) the pumping frequency within a single vessel. Abnormal features, such as highly tortuous or vessels with retrograde lymph flow, were noted.

**Results:** Contraction of lymphatic vessels propelling ICG could be seen in all individuals, with drainage via the anteromedial and anterolateral drainage pathways predominating (observed in 31/32 and 25/32 limbs respectively). The number of lymphatic vessels crossing the anterior ankle was 3.4 ± 1.1 with an average rate of 1 propulsion every 66 seconds (i.e. 0.9 min^-1^) in the vessels investigated. Isolated cases of highly tortuous and refluxing vessels were observed.

**Conclusions:** ICGL is a reliable method for imaging superficial lymphatic vessels in vivo and in real time, helping to establish normal anatomy, vessel numbers and flow characteristics, and so better detect pathological abnormalities.

## Introduction

The lymphatic system has historically been challenging to image which has limited understanding of lymphatics and their contribution to disease. Comprised predominantly of fragile, sub-millimetre lymphatic vessels, filled with colourless lymph fluid, cannulation and the lymphatic delivery of contrast agents for radiographic lymphangiography is technically demanding with potentially severe side effects including pulmonary embolism (1). Lymphatic flow is also slower than blood circulation and erratically pulsatile, making techniques such as Doppler ultrasound not generally applicable to the study of the lymphatics (2).

Indocyanine Green (ICG) lymphography (ICGL) is an imaging technique which offers high spatial and temporal resolution visualisation of superficial lymphatics compared to the more routinely used lymphoscintigraphy (3). Relying on preferential trafficking of the ICG into lymphatic vessels following interstitial injection, ICGL does not require direct lymphatic cannulation (2). Illumination with near infra-red (IR) radiation, and detection of the shifted IR emission, may also be performed with small portable devices providing immediate depiction of the lymphatics. ICGL can therefore be used in the clinic to allow real time and in vivo imaging of the superficial lymphatic vessels with no ionising radiation exposure. Results including the identification of lymphatic pathways, and whether the flow within them is functional or disrupted, can then guide and assess therapies for lymphoedema such as manual lymphatic drainage and lymphovenous anastomosis surgery during treatment (4,5).

ICGL provides valuable novel insights into the functioning of the superficial lymphatic system. Previous studies have used ICGL to study the lymphatics in the extremities of patients with primary and secondary lymphoedema and clearly demonstrate benefits over other investigation techniques (6–10). To recognise pathological abnormalities, one must know what is normal, and yet few reports of lymphatic vessels within normal legs of healthy volunteers have been published. Many studies have imaged the non-oedematous limb of unilateral lymphoedema patients as a comparator to an affected limb, however these limbs could conceal incipient lymphoedema, while studies of cadavers cannot capture true lymphatic physiology (11–13).

In this study, we aim to determine normal parameters for the anatomy and physiology of the superficial lymphatic vessels in the lower limbs of healthy individuals without lymphatic disease using ICGL.

## Methods

### Participants

Healthy volunteers recruited to undergo ICGL in the lower limb at St George’s, University of London (SGUL) between 21/12/2022 and 21/11/2023 had their ICGL data reviewed in this study. Participants included staff members and friends or partners of existing patients, but not their relatives. Prior to enrolment participants were screened for any history of lymphatic disease and, to ensure their safety to undergo ICGL, enquiries regarding any known allergy to ICG, sodium iodide or iodine, were made. Pregnant individuals were excluded.

Ethics approval for this research was granted by the Southeast Coast – Brighton & Sussex Research Ethics Committee, 14/LO/0753.

### ICG Contrast Injection Protocol

To encourage access of ICG into each of the four lymphatic vessel groups of the lower limbs (anteromedial, anterolateral, posterolateral and posteromedial), ICG was administered into 5 locations on each foot: 1^st^ and 4^th^ web spaces, lateral midfoot and lateral and medial rear-foot (Fig 1). Each injection consisted of a 0.1ml mixture comprising 25mg total volume of 1g/L ICG (Verdye™) dissolved into 20 ml water for injection and 5ml 1% lidocaine. ICG was administered through intra-dermal injections manually using a 29G needle and 1 ml syringe. Following injections, participants were then asked to perform foot and toe flexions ten times.

**Figure 1.**
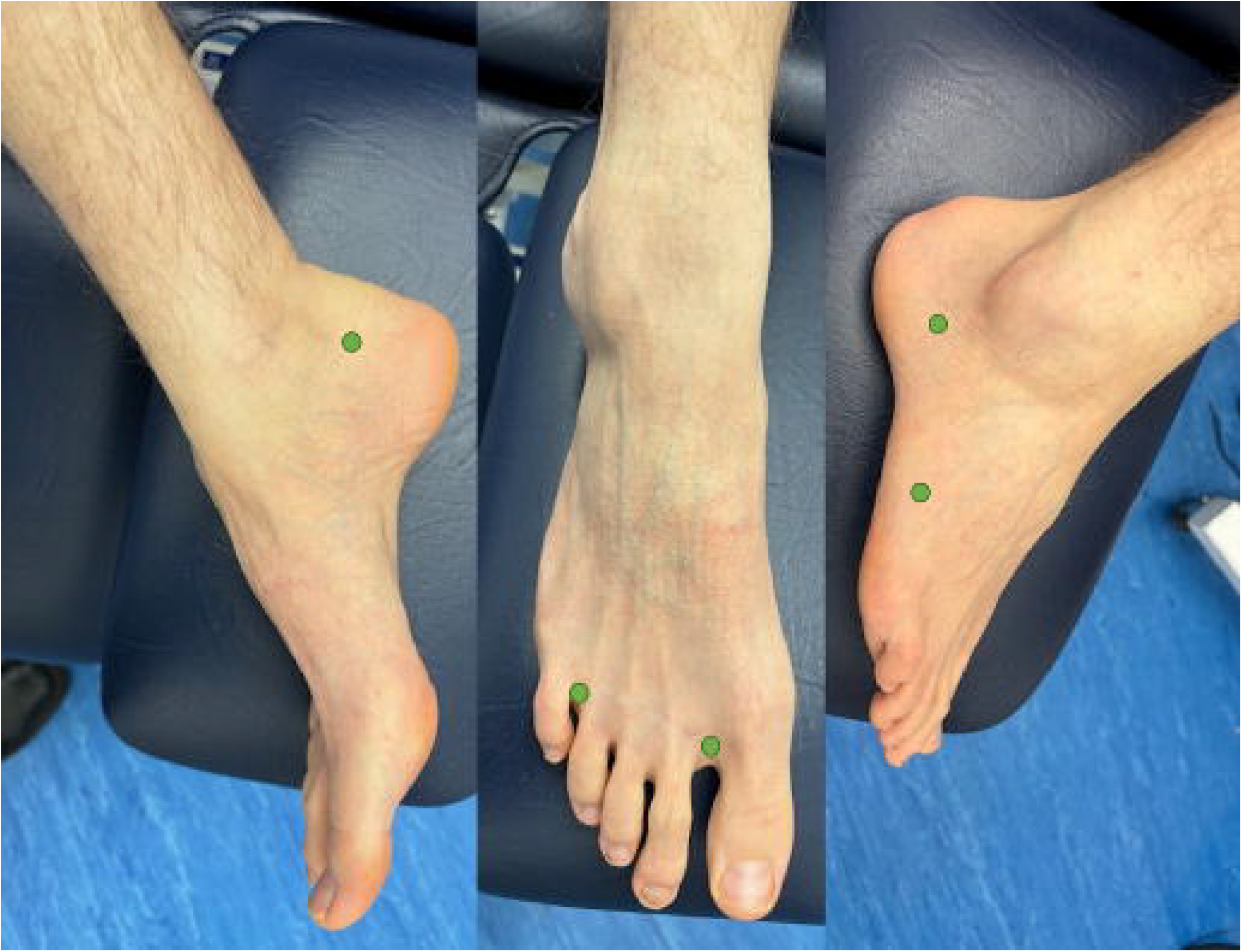
Healthy control foot indicating the five ICG injection points (green dots). Left: medial rear-foot. Middle: 1st and 4th web spaces. Right: lateral midfoot and rear-foot. Adapted from (30).

### Imaging Protocol

ICGL was performed with a Fluobeam 800 (Fluoptics, Grenoble, France), capable of exciting the area under investigation with IR radiation at 750nm and detecting the emitted IR with a single optical head. The optical head was mounted on an articulating arm to improve camera stability whilst imaging.

Imaging consisted of a mobile phase to document the overall pattern of lymphatic enhancement within a limb and videos of between 3-5 minutes focusing on different anatomical regions where lymphatics could be identified. The assessment often included checking the integrity of lymphatic collector one-way valves by the application of manual pressure down the limb. Participants were also routinely asked to stand so that the back of the limb could also be studied.

### Image Analysis

Employing the classification described by Shinaoka et al. (14), lymphatic vessels were categorised based on the path taken in the limb as belonging to one of four groups: anteromedial, posteromedial, anterolateral and posterolateral. Additional imaging measures of interest were: i) the number of lymphatic vessels crossing the anterior ankle (approximately at the level of the talus), ii) the pumping frequency within one of these ankle vessels (i.e. the number of times a vessel emptied during continuous video acquisitions of at least 3 minutes divided by the video duration) and iii) imaging signs considered of an abnormal nature.

Potentially abnormal imaging features included lymphatic vessels that were highly tortuous or directed across, or back down, the limb, the presence of lymph nodes, regions of dermal backflow and lymph stasis (i.e. not being drained from the injection site). The integrity of lymphatic valves was also routinely interrogated by the manual application of pressure distally down the leg. Instances where lymph was manually drainable back down the limb was also considered abnormal.

## Results

Sixteen healthy individuals aged between 20 and 55 years (mean = 38.8; 7 females, 9 males) were recruited and imaged. Lymphatic vessels actively transporting ICG towards the groin were seen in all imaged limbs, and no adverse effects were recorded during or after imaging.

### Lymphatic Anatomy

Contractile linear lymphatic vessels were observed in all 32 limbs of the enrolled participants (Fig 2) and were traceable from injection site up to the groin in 27 of 31 limbs (for a single limb, images of the thigh were unavailable for review). Regions of dermal backflow, lymph stasis and reflux were not observed. In one limb, a vessel looped back on itself and pointed down the leg toward the foot before appearing to merge with another vessel travelling up the leg (Fig 3, supplementary video 1). In two limbs, discontinuous lymphatic vessels were demonstrated, comprising seemingly unconnected vessel fragments, while in a separate limb an unusually tortuous lymphatic vessel fragment was evident (Fig 4).

**Figure 2.**
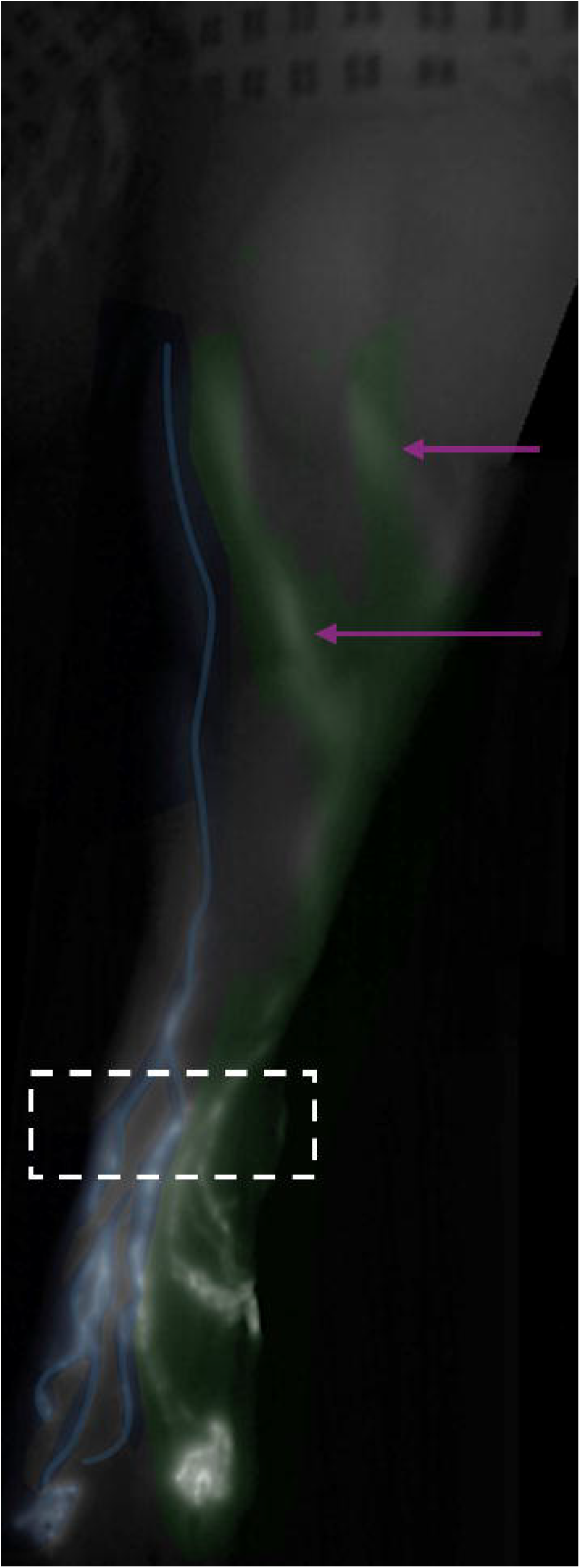
Representative Indocyanine Green Lymphography image of pathways in a healthy female’s left limb demonstrating anteromedial (blue) and anterolateral (green) lymphatic drainage. Four lymphatic vessels were observed passing the anterior aspect of the ankle (hashed box arrow). Note that some lateral vessels moving medially just below the level of the knee (arrows) and the injection sites in foot visible at the bottom of the image.

**Figure 3.**
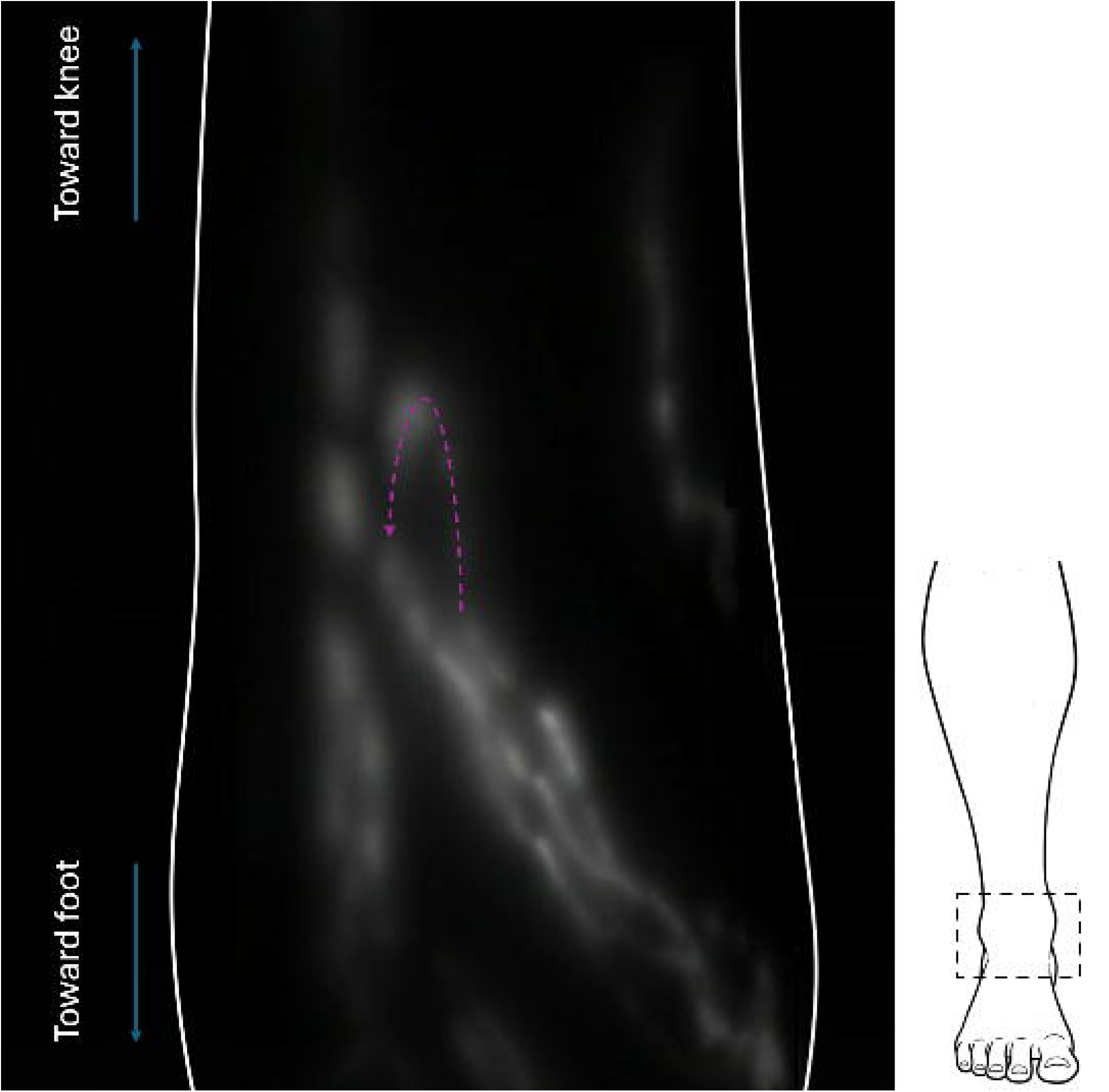
Lymphatic3 vessel looping back on itself (purple arrow showing the direction of flow) and pointing down toward the foot at the anterior ankle of a healthy female’s left foot (limb outline shown in white) before appearing to merge with another vessel travelling up the leg (see supplementary video 1. The schematic on the right shows the approximate location of the imaged region (hashed box).

**Figure 4.**
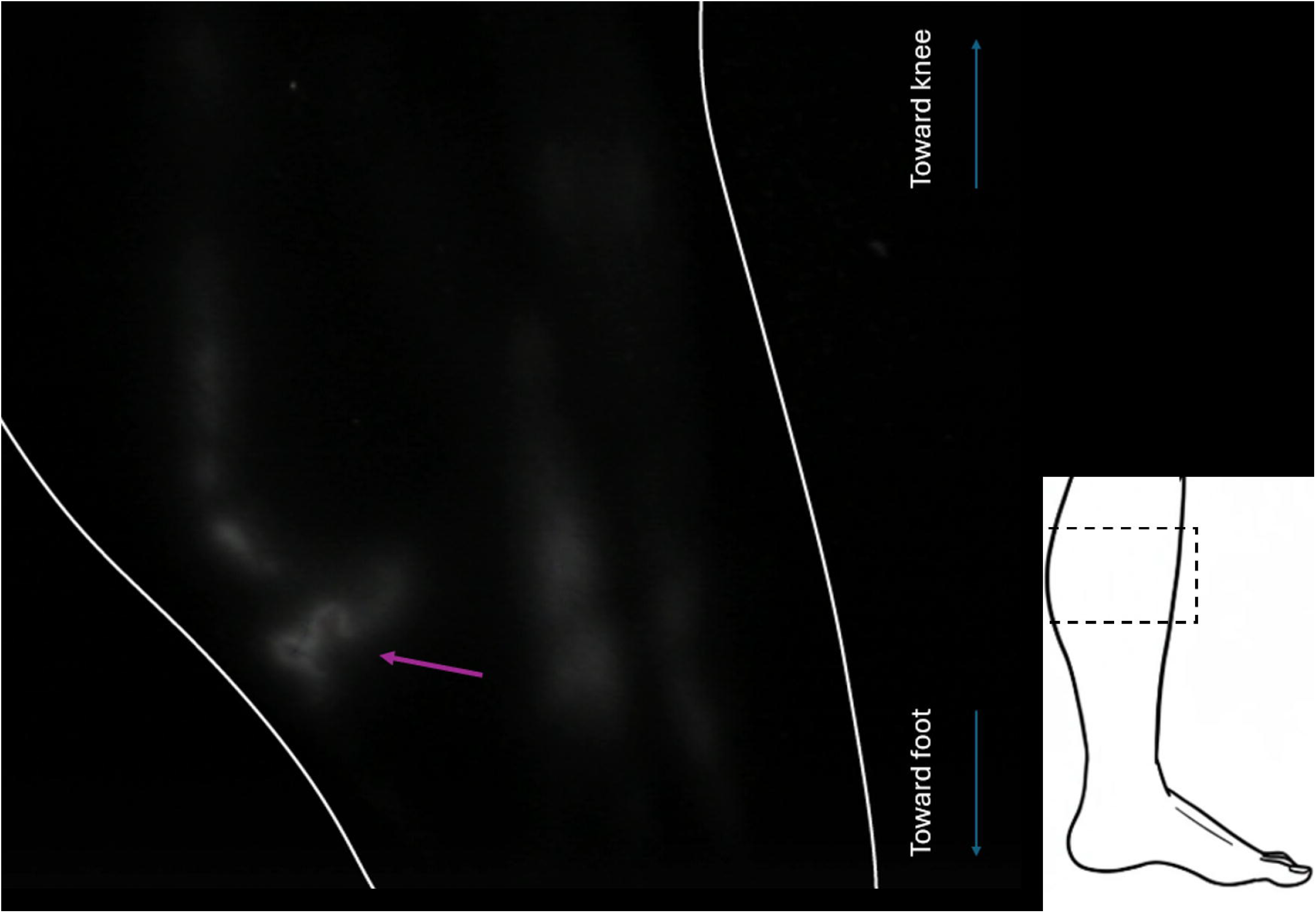
Small tortuous vessel segment located at the medial calf in a healthy individual (the same individual as shown in Figure 3.). Lymphatic vessels with a curvilinear nature were commonly observed as they travelled up the limb, but such tightly tortuous vessels (arrow) were identified only in one limb (limb outline shown in white). The schematic on the right shows the approximate location of the imaged region at the medial aspect of the leg (hashed box).

Anteromedial and anterolateral pathways were observed in 31/32 and 25/32 limbs respectively (Fig 2), and all limbs had at least one, and at most 5, lymphatic vessels observed passing the anterior aspect of the ankle (mean ± standard deviation = 3.4 ± 1.1; mode = 4). In 18 limbs, posteromedial enhancement was noted, while in only 9 was posterolateral. In all but three cases (29 of 32 limbs) the posterior aspect of the leg (calf) was imaged. Of the 29, a popliteal node was detected in 12 (Fig 5) and popliteal vessels in 13. For 6 limbs, popliteal lymph nodes and vessels were observed as isolated structures within the popliteal fossa without corresponding posterolateral lymphatic pathways being observed.

**Figure 5.**
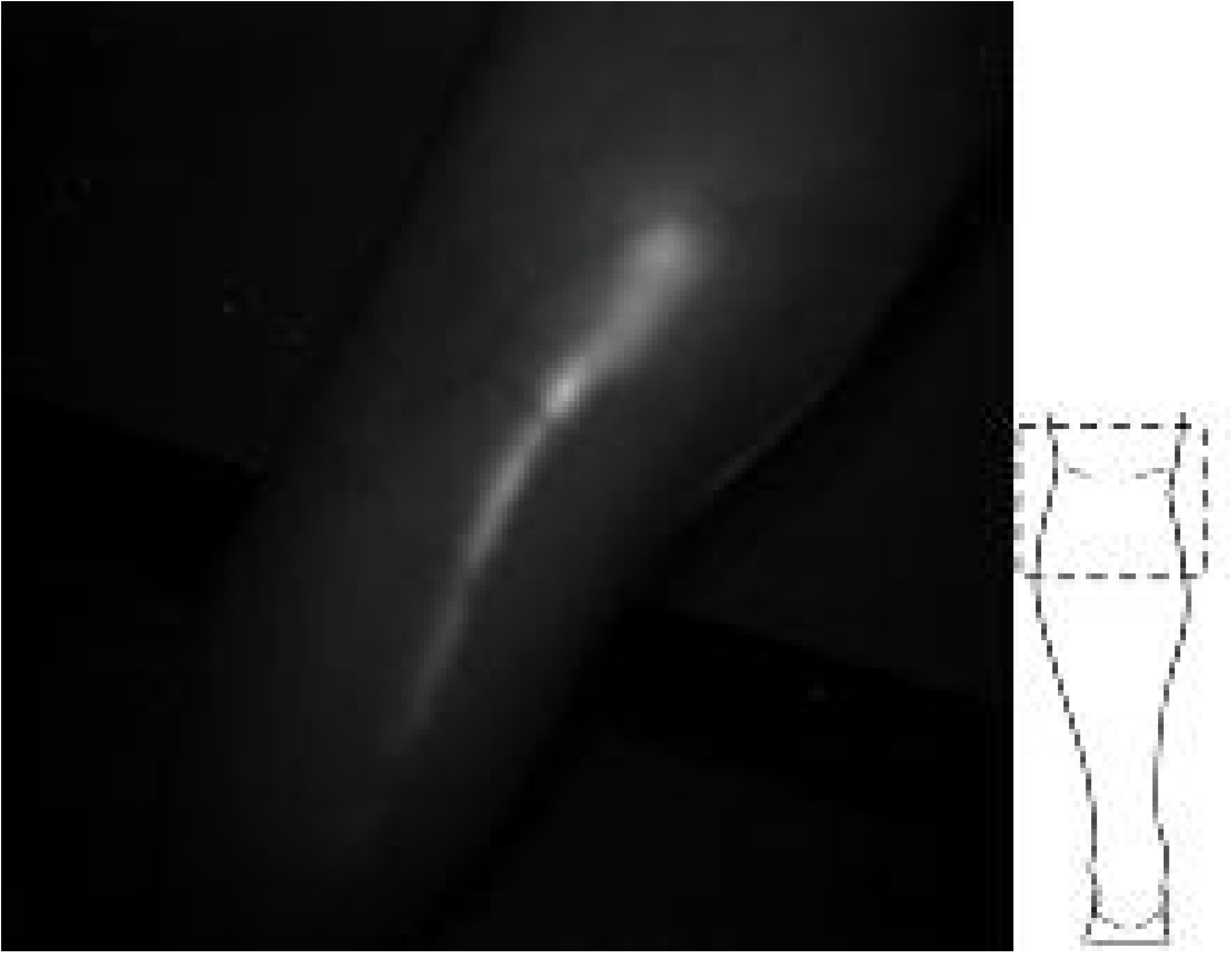
Posterolateral lymphatic vessel transporting ICG from the rear leg to a lymph node (arrow) in the popliteal fossa in a male volunteer. No efferent vessels from the node into the thigh (upper portion of the image) was demonstrated. The schematic of the rear of the limb shows the approximate location of the imaged region (hashed box).

### Lymphatic Pumping

An average pumping frequency of 0.9 ± 0.4 min^-1^ (range = 0 – 2min^-1^) was estimated in anterior ankle lymphatics from videos lasting between 180 – 250 seconds with between 0-7 (mode = 2) pumping events observed.

Of 20/32 limbs investigated, in only one participant was ICG manually drainable back down the limb (both left and right) suggesting valve incompetence (supplementary video 2).

## Discussion

Indocyanine green (ICG) lymphography has emerged as an important technique for studying the superficial lymphatic system. It is particularly useful for studying vessels too small to observe with Magnetic Resonance Imaging (MRI) or lymphoscintigraphy (LS) and has been deployed in previously difficult regions to image such as the genitals(15–17). Its advantages are real time and in vivo visualisation of lymphatic transport. ICGL’s clinical applicability is enhanced further by its minimally invasive and non-ionising nature. However, like any tracer-based technique, it is limited by possible contraindications to injection of a foreign agent and pain at administration. The poor penetrance of the infrared light also makes imaging vessels below around 2cm in the skin impossible and the acquisition of image data is highly user dependent.

Additionally, our recent review highlighted the need to develop a consensus protocol for the performance and interpretation of ICGL studies, and the presentation of consistent participant and imaging features (10). Standardised and validated protocols are of great importance for improving diagnostic accuracy, the direct comparison of study findings, and the translation of ICGL from a research focussed tool to something more clinical useful (9,18).

In this study, we focused exclusively on the imaging of healthy subjects, attempting to establish normal lymphatic anatomy and function. Imaging healthy controls, and not the contralateral limb of unilaterally affected lymphoedema patients where subclinical disease may be present (12,19), is essential for establishing what normal lymphatic anatomical structures and functioning look like. Only then can one determine what is abnormal and what represents pathology, enabling improved disease detection and identification of disease-specific changes.

The sites for administration of ICG were chosen to induce drainage via each of the four pathways described by Shinaoka et al. (20). In most limbs, drainage via the anteromedial and anterolateral pathways was identified. In the single limb in which anteromedial vessels were not seen, anterolateral vessels were and so all limbs demonstrated vessels crossing the anterior ankle. This likely represents normal anatomical variation within individuals, similar to what is seen with the veins (21).

Vessels following the posterolateral pathway were identified infrequently (9 limbs). Reduced visibility of lymphatic pathways in lymphoedema patients with ICGL can result from increased skin thickness and adipose tissue deposition. Given that all participants in this study were lymphoedema free, we suspect that the lower incidence of posterior lymphatic visualisation results from the vessels simply residing deeper within the limb and the limited penetration depth of ICG lymphography (around 2cm). Despite the poor penetrance, lymphatic vessels could generally be detected all the way to the groin.

Within the thigh and groin, substantial scattering of the emitted light was evident, and the vessels faint and blurred, likely a result of the deeper course of the lymphatic vessels in the thigh compared to the lower leg. Functional assessment of the lymphatics in the lower limb is therefore most likely to be most sensitive in the leg (below the knee) as opposed to the thigh, and justifies our choice to assess valve competency, vessel count and pumping frequency in the ankle and leg region.

Although infrequent, some features that may be indicative of lymphatic dysfunction or abnormality were observed in this healthy control cohort, including unusually tortuous vessel segments, lymphatic flow oriented down a limb, and regions where ICG could be manually pushed back down the limb, suggestive of valve incompetence. None of our participants in this study reported a history of lymphatic disease, however we cannot rule out the presence of sub-clinical lymphatic abnormalities or previous undisclosed injury being the cause of these findings. Isolated cases such as these may also simply be natural variations in these individuals’ lymphatic systems, particularly when compared to the widespread abnormalities described in patient studies (9,22,23), or, in the case of discontinuous vessel fragments, due to limitations in the penetration of the ICG imaging technique.

The presence of popliteal lymph nodes in lymphoscintigrams has been reported as suggesting pathological rerouting of the radionuclide via the deep lymphatics(24,25). However, popliteal nodes were regularly observed in our healthy cohort. The common visualisation of these nodes is likely due to our injecting of ICG into a location in the foot which has been shown to elicit drainage via the posterolateral lymphatic pathway and to popliteal lymph nodes. Clinicians need to be cognizant of the injection sites for any dyes or tracers used in imaging studies therefore and whether popliteal enhancement could be expected to occur even in healthy individuals.

Lymphatic aplasia, errors with lymphatic pumping and valve incompetence have all been recognised as mechanisms for causing lymphoedema. Measurement of lymphatic contractile frequency and the speed of ICG bolus movement in lymphatic vessels have previously been attempted in the arms of healthy controls (26,27), and the arrival time of ICG at the groin of lower limb lymphoedema patients following ICG injections in the foot (28). This shows the potential use of ICGL as a tool for quantifying lymphatic transport, but to our knowledge no report has established similar measurements for the lower limbs of healthy individuals. Establishing normal ranges for quantitative parameters such as these would be beneficial for improving the detection of lymphatic abnormalities, particularly those subtle changes which are not yet manifesting a clinical effect (3).

Investigation of the anterior ankle enabled observation of linear vessels in all individuals, with 1-5 vessels visualised across all limbs. An average pumping rate of 0.9 min^-1^ was measured in the single vessel interrogated in each limb. Only a single vessel was interrogated in each limb to reduce measurement error in less clearly resolved vessels, and due to the time taken to make these observations. Some processing pipelines have been proposed to improve the efficiency and accuracy of monitoring lymphatic contractions with ICGL (26,29), but do not appear to be in widespread use. Future research exploring the deployment of these tools may increase their adoption and further the use of ICGL for quantitative lymphatic assessment.

## Conclusion

Healthy superficial lymphatic anatomy and functioning can reliably be studied in vivo and in real time with ICGL. In this study of individuals with no history of lymphatic disease, consistent findings were established with contractile superficial lymphatic vessels observed in all imaged legs. We have established that not all four lymphatic vessel groups of the lower limbs (anteromedial, anterolateral, posterolateral and posteromedial) will be observed in all cases, even with ICG injection sites chosen to access them. The lymphatic vessels which were observed were normally linear, with an average of 3 seen crossing the anterior ankle. Valve incompetency can be tested via the application of pressure down the limb and, along with tortuous lymphatic vessels, is generally absent in healthy legs. Tortuous vessels and valve reflux should therefore be considered abnormal.

Only by establishing normal parameters for anatomy, vessel numbers and direction of flow, can real pathological abnormalities be determined. ICGL is easy-to-use in the clinic and could be a valuable diagnostic tool if objective outcome measures, as developed in this study, are available. This would not only aid diagnosis of lymphoedema but also help determine phenotypes for genetic studies. ICGL could prove useful for the diagnosis of lymphatic abnormalities in previously understudied regions, such as the face, breast and genital regions, and improve patient management by demonstrating measured responses to therapeutic interventions.

## Data Availability

All data produced in the present study are available upon reasonable request to the authors

## Figure Captions

Supplementary Video 1. In a single limb, a vessel looped back on itself and pointed down the leg before appearing to merge with another vessel travelling up the leg. This was evidently the direction of flow and not reflux for this vessel as multiple pumping events were clearly observed in this vessel over the course of four minutes (30s video shown for brevity). Interestingly, this induvial also demonstrated a small tortuous lymphatic vessel segment which is shown in Figure 4. of the main manuscript. Note that the orientation of this video matches Figure 3. in the main manuscript, with the superior aspect of the limb at the top of the frame and the inferior at the bottom.

Supplementary Video 2. Video of a female volunteer’s leg in which it is evident that lymph containing ICG can be forced back down the limb (toward the top of the frame) by applying pressure to the limb. This suggests incompetent valves within this individual’s lower limb lymphatics. Once the hand is removed, the lymphatic fluid rapidly flows back up the limb. A similar finding was noted for both lower limbs.

## References

1. Jackson RJA, Brit BAOMY. Complications of Lymphography. Vol. 1. 1966.

2. Lee B boong, Rockson SG. Lymphedema: A Concise Compendium of Theory and Practice [Internet]. Second Edi. Lee BB, Rockson SG, Bergan J, editors. Cham: Springer International Publishing; 2018. Available from: http://link.springer.com/10.1007/978-3-319-52423-8

3. Figueroa BA, Lammers JD, Al-Malak M, Pandey S, Chen WF. Lymphoscintigraphy versus Indocyanine Green Lymphography—Which Should Be the Gold Standard for Lymphedema Imaging? Lymphatics. 2023 May 5;1(1):25–33.

4. Ogata F, Narushima M, Mihara M, Azuma R, Morimoto Y, Koshima I. Intraoperative lymphography using indocyanine green dye for near-infrared fluorescence labeling in lymphedema. Ann Plast Surg. 2007 Aug;59(2):180–4.

5. Koelmeyer LA, Thompson BM, Mackie H, Blackwell R, Heydon-White A, Moloney E, et al. Personalizing conservative lymphedema management using indocyanine green-guided manual lymphatic drainage. Lymphat Res Biol. 2021 Feb 1;19(1):56–65.

6. Burnier P, Niddam J, Bosc R, Hersant B, Meningaud JP. Indocyanine green applications in plastic surgery: A review of the literature. Journal of Plastic, Reconstructive and Aesthetic Surgery [Internet]. 2017;70(6):814–27. Available from: http://www.elsevier.com

7. Kato M, Watanabe S, Iida T, Watanabe A. Flow Pattern Classification in Lymphatic Malformations by Indocyanine Green Lymphography. Plast Reconstr Surg [Internet]. 2019;143(3):558E–564E. Available from: http://ovidsp.ovid.com/ovidweb.cgi?T=JS&PAGE=reference&D=emed20&NEWS=N&AN=626595069

8. Chao AH, Schulz SA, Povoski SP. The application of indocyanine green (ICG) and near-infrared (NIR) fluorescence imaging for assessment of the lymphatic system in reconstructive lymphaticovenular anastomosis surgery. Expert Rev Med Devices [Internet]. 2021;18(4):367–74. Available from: http://ovidsp.ovid.com/ovidweb.cgi?T=JS&PAGE=reference&D=med19&NEWS=N&AN=33686906

9. Suami H, Thompson B, Mackie H, Blackwell R, Heydon-White A, Blake FT, et al. A new indocyanine green fluorescence lymphography protocol for diagnostic assessment of lower limb lymphoedema. Journal of Plastic, Reconstructive and Aesthetic Surgery [Internet]. 2022;75(11):3946–55. Available from: 10.1016/j.bjps.2022.08.017

10. Brezgyte G, Mills M, van Zanten M, Gordon K, Mortimer PS, Ostergaard P. A systematic review of indocyanine green lymphography imaging for the diagnosis of primary lymphoedema. British Journal of Radiology [Internet]. 2025 Jan 21; Available from: https://academic.oup.com/bjr/advance-article/doi/10.1093/bjr/tqaf006/7965899

11. Shinaoka A, Koshimune S, Yamada K, Kumagishi K, Suami H, Kimata Y, et al. A Fresh Cadaver Study on Indocyanine Green Fluorescence Lymphography: A New Whole-Body Imaging Technique for Investigating the Superficial Lymphatics. Plast Reconstr Surg. 2018;141(5):1161–4.

12. de Almeida CA, Lins EM, Brandão SCS, Ferraz ÁAB, Pinto FCM, de Barros Marques SR. Lymphoscintigraphic abnormalities in the contralateral lower limbs of patients with unilateral lymphedema. J Vasc Surg Venous Lymphat Disord. 2017 May 1;5(3):363–9.

13. Burnand KM, Glass DM, Mortimer PS, Peters AM. Lymphatic dysfunction in the apparently clinically normal contralateral limbs of patients with unilateral lower limb swelling. Clin Nucl Med. 2012;37(1):9–13.

14. Shinaoka A, Koshimune S, Suami H, Yamada K, Kumagishi K, Boyages J, et al. Lower-limb lymphatic drainage pathways and lymph nodes: A CT lymphangiography cadaver study. Radiology. 2020;294(1):223–9.

15. Frojo G, Castro O, Tadisina KK, Xu KY. Lymphovenous Bypass Using Indocyanine Green Mapping for Successful Treatment of Penile and Scrotal Lymphedema. Plast Reconstr Surg Glob Open. 2020;8(7).

16. Yuan Y, Li F, Zhou Y, Li S, Cao Y, Liu M, et al. Lymphatic Pathways on Indocyanine Green Lymphography in Patients with Labia Minora Hypertrophy. Plast Reconstr Surg. 2024 Sep 1;154(3):665–71.

17. Hara H, Mihara M. Indocyanine Green Lymphographic and Lymphoscintigraphic Findings in Genital Lymphedema-Genital Pathway Score. Lymphat Res Biol. 2017 Dec 1;15(4):356–9.

18. Suami H, Heydon-White A, Mackie H, Czerniec S, Koelmeyer L, Boyages J. A new indocyanine green fluorescence lymphography protocol for identification of the lymphatic drainage pathway for patients with breast cancer-related lymphoedema. BMC Cancer. 2019 Oct 22;19(1).

19. Aldrich MB, Guilliod R, Fife CE, Maus EA, Smith L, Rasmussen JC, et al. Lymphatic abnormalities in the normal contralateral arms of subjects with breast cancer-related lymphedema as assessed by near-infrared fluorescent imaging. 2012.

20. Shinaoka A, Koshimune S, Suami H, Yamada K, Kumagishi K, Boyages J, et al. Lower-limb lymphatic drainage pathways and lymph nodes: A CT lymphangiography cadaver study. Radiology. 2020;294(1):223–9.

21. B CocKIIrm BF, St Thomas F. ABNORMALITIES OF THE DEEP VEINS OF THE LEG.

22. Shinaoka A, Kamiyama K, Yamada K, Kimata Y. A new severity classification of lower limb secondary lymphedema based on lymphatic pathway defects in an indocyanine green fluorescent lymphography study. Sci Rep [Internet]. 2022;12(1):1–11. Available from: 10.1038/s41598-021-03637-6

23. Tashiro K, Yamashita S, Saito T, Iida T, Koshima I. Proximal and distal patterns: Different spreading patterns of indocyanine green lymphography in secondary lower extremity lymphedema. Journal of Plastic, Reconstructive and Aesthetic Surgery. 2016 Mar 1;69(3):368–75.

24. Burnand KM, Glass DM, Sundaraiya S, Mortimer PS, Peters AM. Popliteal node visualization during standard pedal lymphoscintigraphy for a swollen limb indicates impaired lymph drainage. American Journal of Roentgenology. 2011 Dec;197(6):1443–8.

25. Kandeel AAS, Younes J, Zaher AM. Significance of popliteal lymph nodes visualization during radionuclide lymphoscintigraphy for lower limb lymphedema. Indian Journal of Nuclear Medicine. 2013 Jul;28(3):134–7.

26. Gray RJ, Worsley PR, Voegeli D, Bader DL. Monitoring contractile dermal lymphatic activity following uniaxial mechanical loading. Med Eng Phys. 2016;38(9):895–903.

27. Lopera C, Worsley PR, Bader DL, Fenlon D. Investigating the Short-Term Effects of Manual Lymphatic Drainage and Compression Garment Therapies on Lymphatic Function Using Near-Infrared Imaging. Lymphat Res Biol. 2017 Sep 1;15(3):235–40.

28. Yoshida S, Koshima I, Imai H, Sasaki A, Fujioka Y, Nagamatsu S, et al. Indocyanine green lymphography findings in older patients with lower limb lymphedema. J Vasc Surg Venous Lymphat Disord [Internet]. 2020;8(2):251–8. Available from: http://www.elsevier.com/journals/journal-of-vascular-surgery-venous-and-lymphatic-disorders/2213-333X

29. Granoff MD, Johnson AR, Lee BT, Padera TP, Bouta EM, Singhal D. A Novel Approach to Quantifying Lymphatic Contractility during Indocyanine Green Lymphangiography. Plast Reconstr Surg [Internet]. 2019;144(5):1197–201. Available from: http://ovidsp.ovid.com/ovidweb.cgi?T=JS&PAGE=reference&D=emed20&NEWS=N&AN=629767044

30. Brezgyte G, Mills M, Heenan S, Gordon K. Indocyanine green lymphography identifies lymphatic structures in patients with Milroy’s disease who demonstrate functional aplasia on lymphoscintigraphy. BMJ Case Rep [Internet]. 2024 Nov 21;17(11):e261787. Available from: https://casereports.bmj.com/lookup/doi/10.1136/bcr-2024-261787

